# Transitions across Kidney Disease Stages and Association with New-onset Heart Failure: a Population-Based study of 8569 Individuals and 12.6 Years of Follow-up

**DOI:** 10.1101/2022.12.31.22284047

**Authors:** Yuntao Chen, Stephan JL Bakker, Navin Suthahar, Laura M.G. Meems, Dion Groothof, Dirk J. van Veldhuisen, Wiek H. van Gilst, Rudolf A de Boer, Ron T Gansevoort, Gerton Lunter, Hans L. Hillege, Douwe Postmus

## Abstract

**Background:** Limited data exist on transitions across Kidney Disease Improving Global Outcomes (KDIGO) risk stages and association with new-onset heart failure (HF) in the general population.

**Methods:** Data were extacted from the first four clinical examinations of the PREVEND study. Participants were categorized into eGFR and albuminuria risk stages according to KDIGO Guideline. Multi-state model was fitted to estimate the transition rates across consecutive KDIGO risk stages and the association with new-onset HF.

**Results:** 8569 participants were included at baseline. During a median follow-up of 12.6 years, 373 participants were diagnosed with new-onset HF and 710 participants had died before new-onset HF. The transition rate from low risk to intermediate risk stage did not differ between women and men; it increased from 0 to 10 transitions per 100 person-years from age 50 to 75 years. The transition rate from intermediate risk to high or very high risk stage increased exponentially with age, with women having lower rates (HR: 0.50, 95%CI: 0.34-0.76). Younger subjects and women were more likely to make a backward transition from intermediate risk to low risk stage. More advanced KDIGO risk stages were associated with increased risk of new-onset HF, and the risk association attenuates with age on the relative scale but strengthens with age on the absolute scale.

**Conclusions:** Slowing the progression of KDIGO risk stages may result in an important reduction in the incidence of new-onset heart failure in this population of middle-aged subjects.

## Introduction

It is well established that individuals with chronic kidney disease (CKD) are at increased risk of developing heart failure (HF)^1,2^. Moreover, evidence suggests that this association is modified by age, with stronger relative effects in young subjects and weaker relative effects in elderly subjects^3,4^. What further complicates studying the cardiorenal syndrome is that age not only acts as an effect-modifier, but also a risk factor for both decline of renal function^5^ and development of HF. This already complex effect of age may be further complicated by sex, because it has been recognized that behavioural and biological differences between the sexes lead to differences in the time course of both CKD and HF^5–7^. Finally, the association between renal function and the development of HF is dependent on the measure of renal function used^8,9^. A large meta-analysis found that the risk of developing HF increased linearly with urinary albumin excretion (UAE), while this relationship was found to be non-linear for estimated glomerular filtration rate (eGFR), with little change in risk when eGFR was higher than 75 mL/min per 1.73m^2^ and linearly increasing risk with decreasing eGFR below this threshold^10^. In population-based cohorts with mostly healthy subjects, UAE is therefore likely to be a stronger predictor for new-onset HF than eGFR^9^. The Kidney Disease Improving Global Outcomes (KDIGO) guideline used eGFR and UAE to categorize a population into low, intermediate, high, or very high risk on the basis of evidence from the Chronic Kidney Disease Prognosis Consortium (CKD-PC)^11^.

To better understand the complex interplay between age, sex, kidney function status, and the development of HF, it is essential to consider a subject’s entire disease course from the initial kidney function stage at baseline to the development of HF. By modelling the evolution of a subject’s kidney function status as transitions between consecutive KDIGO risk stages defined in terms of eGFR and UAE, the use of multi-state modelling allows for a more detailed representation of how a subject’s kidney function changes over time and how these longitudinal changes can be linked to later adverse outcomes such as the development of HF. While multi-state modelling has previously been applied to model the progression of a range of diseases, including dementia^12^ and age-related macular degeneration^13^, this approach has not been applied in studying the development of HF in the cardiorenal syndrome.

By fitting a multi-state model to the longitudinal data from the PREVEND study, a prospective cohort study performed in the city of Groningen in the Netherlands, the purpose of this research was to estimate age- and sex-specific transition rates between consecutive KDIGO risk stages before new-onset HF. Besides, we sought to estimate age-, sex-specific association of KDIGO risk stages with new-onset HF taking into account dynamic change and potential misclassification in kidney measures.

## Methods

### Study Population

The present study was performed using data from the Prevention of REnal and Vascular ENd-stage Disease (PREVEND) study, a prospective cohort study that was set up to evaluate the association of increased UAE with cardiovascular and kidney disease in apparently healthy individuals. Details of the study protocol have been published elsewhere^14^. In brief, all inhabitants of the city of Groningen between the ages of 28 and 75 years old (N = 85,421) were invited to answer a postal questionnaire and to provide an early morning urine sample. The response rate was 47.8% (N = 40,856), and urinary albumin and creatinine levels were measured in these respondents. Pregnant women and those with prevalent type 1 diabetes mellitus were excluded. All remaining subjects with a UAE of ≥ 10 mg/L (N = 6000) and a randomly selected control group with UAE < 10 mg/L (N = 2592) underwent further investigation and constituted the baseline PREVEND cohort. The PREVEND study was approved by the institutional medical ethics committee and conducted in accordance with the Declaration of Helsinki. All subjects provided written informed consent.

### Measures

#### Kidney function measures

We used data collected at the first four clinical examinations of the PREVEND study (1997-1998, 2001-2003, 2003-2006, 2006-2008) to extract information on UAE and eGFR. UAE was calculated as the average UAE in two consecutive 24-hour urine collections. For the primary analyses, eGFR was calculated from serum creatinine standardized to isotope dilution mass spectrometry. To examine for potential confounding by muscle mass and/or diet^15,16^, a sensitivity analysis was conducted using eGFR calculated from IFCC-calibrated serum cystatin C^17^. The Chronic Kidney Disease Epidemiology Collaboration (CKD-EPI) equation^18^ was used to calculate both the serum creatinine-based and serum cystatin C-based eGFR. Serum creatinine and serum cystatin C were determined in a single run to avoid laboratory day-to-day variation. By combining the two kidney function measures, namely UAE and eGFR, participants were categorized into three risk categories according to the [2012 KDIGO Clinical Practice Guideline for the Evaluation and Management of CKD](https://kdigo.org/guidelines/ckd-evaluation-and-management/)^11^: low risk stage (eGFR > 60 mL/min/1.73 m^2^ and UAE < 30 mg/24 hours), moderately increased risk stage (eGFR > 60 mL/min/1.73 m^2^ and UAE 30-300 mg/24 hours; eGFR 45-59 mL/min/1.73 m^2^ and UAE < 30 mg/24 hours) and high to very high risk stage (eGFR > 60 mL/min/1.73 m^2^ and UAE > 300 mg/24 hours; eGFR 45-59 mL/min/1.73 m^2^ and UAE ≥ 30 mg/24 hours; eGFR < 45 mL/min/1.73 m^2^).

#### New-onset heart failure

The outcome of interest was new-onset HF. Details about how new-onset HF cases were collected and defined are described elsewhere^19^. In brief, an endpoint adjudication committee consisting of seven independent experts in the field of HF evaluated all cases suspected for the diagnosis of new-onset HF. Two experts evaluated each case by reviewing anonymized clinical charts, hospitalization, and physician office records, based on the HF guidelines of the European Society of Cardiology^20^. In consensus, patients were defined as ‘definite new-onset heart failure’; otherwise, the committee made a joint decision. The follow-up for the present study was defined as the time from the baseline visit to the date of new-onset HF, the date of death, or 31 December 2010, whichever came first. Information on the date of death was obtained from Statistics Netherlands. Subjects were censored at the date they moved to an unknown destination or at the last date of the follow-up (31 December 2010).

#### Covariates

Covariates assessed at baseline included hypertension, hypercholesterolaemia, type 2 diabetes, and smoking. Systolic and diastolic blood pressure were calculated as the mean of the last two measurements of the two visits, using an automatic Dinamap XL Model 9300 series device. Hypertension was defined as systolic blood pressure > 140 mmHg, diastolic blood pressure > 90 mmHg, or self-reported use of antihypertension medication.

Hypercholesterolaemia was defined as total serum cholesterol > 6.5 mmol/L or a serum cholesterol ≥ 5.0 mmol/L if a history of myocardial infarction was present or when lipid-lowering medication was used. Type 2 diabetes was defined as a fasting plasma glucose > 7.0 mmol/L, a non-fasting plasma glucose > 11.1 mmol/L or the use of anti-diabetic medication. Smoking was defined as current nicotine use or quitting smoking within the previous year.

### Statistical analyses

To estimate the transition rates between consecutive KDIGO risk stages before incident HF, we fitted a multi-state model with the three KDIGO risk stages as transient states and new-onset HF and death before HF as absorbing states. The model included all forward and backward transitions between consecutive KDIGO risk stages and each risk stage to new-onset HF and death except the backward transition from high or very high risk stage to intermediate risk stage (Figure 1). We did not include this backward transition because it was rare and likely to be due to misclassification. This was evidenced by comparing the full model and the model without this backward transition based on the likelihood ratio test (p=0.70). To account for possible misclassification in the observed risk stages due to variation in eGFR and/or UAE measurements, the observed risk stages were allowed to deviate from the true underlying stages according to a set of misclassification probabilities that were also estimated from the data.

**Figure 1.**
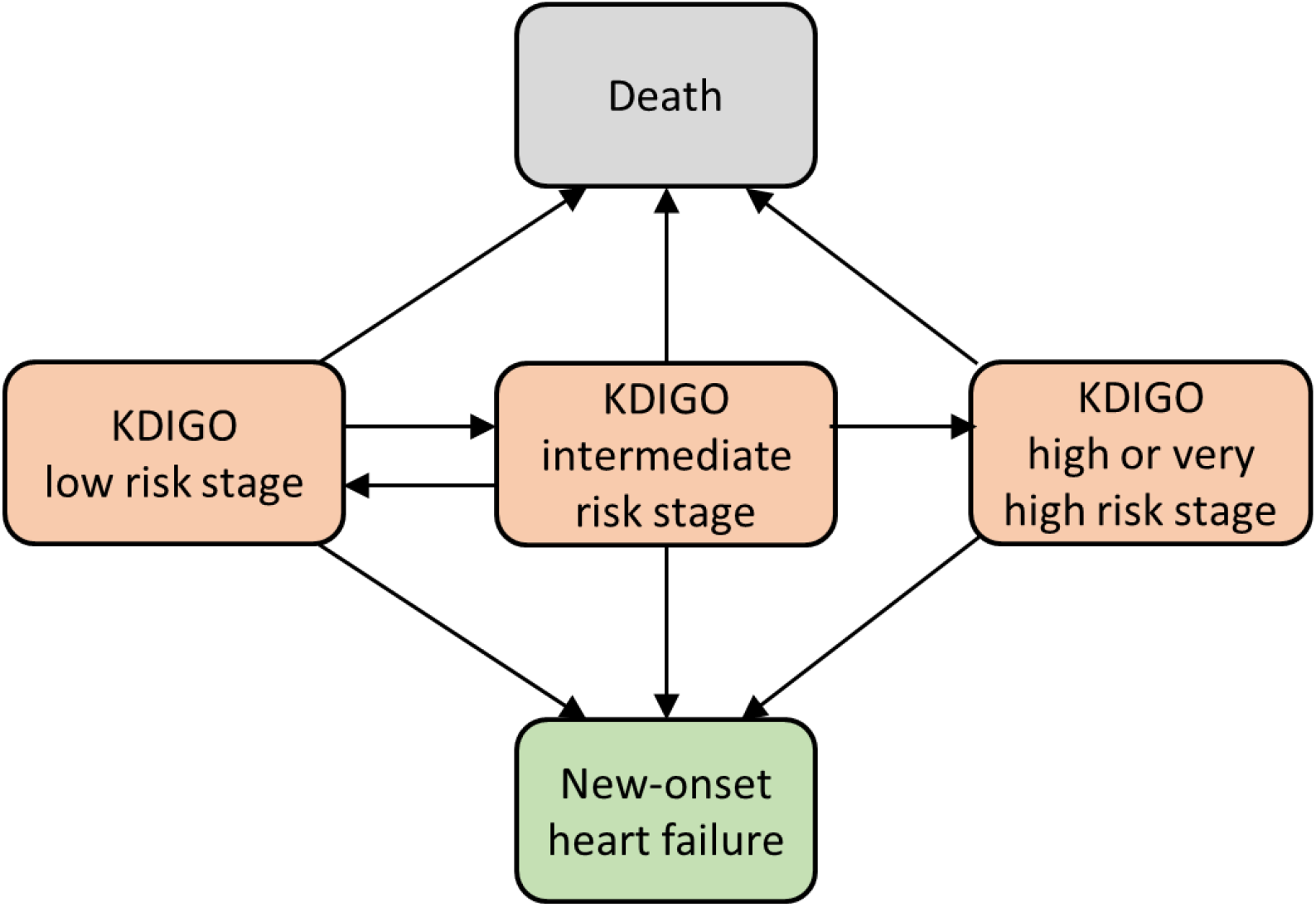
The multi-state Markov model. Each node denotes one of the five states in the model. The new-onset heart failure state was treated as an absorbing state, with death as a competing absorbing state. The three intermediate kidney disease stages were defined according to the 2012 KDIGO Clinical Practice Guideline for the Evaluation and Management of CKD. KIDIGO Low risk stage includes G1A1, G2A1; KDIGO intermediate risk stage includes G1A2, G2A2, G3aA1; KDIGO high or very high risk stage includes G3aA2, G3b, any G4, G5, or A3.

The transition rates were assumed to have the following form:

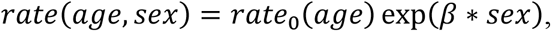

where *age* was used as the time scale and *sex* was incorporated as a covariate (dummy coded as 1 if the *sex* is woman, and 0 otherwise). The baseline rate *rate*_0_(*age*) was initially parameterized as a log-linear function of age, i.e., *rate*_0_ = exp (*α*_0_ + *α*_1_*age*). Likelihood ratio tests were subsequently conducted to determine whether relaxing the log-linearity assumptions via restricted cubic splines improved the model fit. We also used likelihood ratio tests to examine whether the effect of age (i.e., *α*_1_) was the same across the different transitions for which the log-linearity assumption was found to hold. Finally, we used likelihood ratio tests to examine whether the effect of sex (i.e., *β*) was the same across the different transitions to HF and death, respectively.

To demonstrate how the estimated transition rates could be used to explain the observed difference in KDIGO stage prevalence, we used the fitted model to predict the evolution of the KDIGO stage prevalance in a homogeneous population of healthy individuals (low risk stage) aged 50 as this population ages from 50 to 90 years.

Using the estimated transition rates to HF, we calculated age- and sex-specific rate ratios (RRs) for the relative effect of KDIGO risk stage on the risk of new-onset HF. Absolute rate differences (RDs) were also calculated, and their standard errors were obtained using a parametric bootstrap approach^21^. Adjusted RRs and RDs were calculated based on model further adjusting for smoking, hypertension, hypercholesterolaemia and diabetes.

Goodness of fit was assessed by comparing the cumulative incidence curves of new-onset HF and death estimated using the multi-state model with those estimated using the non-parametric Aalen-Johansen estimator. This assessment was performed in the total population as well as in subpopulations divided by sex (men, women), tertiles of baseline age (<42, 42-54, ≥55 years), and KDIGO risk stage at baseline. All analyses were conducted in R, version 3.6.3, using the MSM package^22^ to fit the multi-state model.

## Results

At baseline, 8569 participants were included in the PREVEND study population, with 6,838 (79.8%) in low risk stage, 1,279 (14.9%) in intermediate risk stage, and 303 (3.5%) in high or very high risk stage. Participants in the higher risk stage were older, more likely to be men, and more likely to have hypertension or type 2 diabetes (Table 1).

**Table 1.**
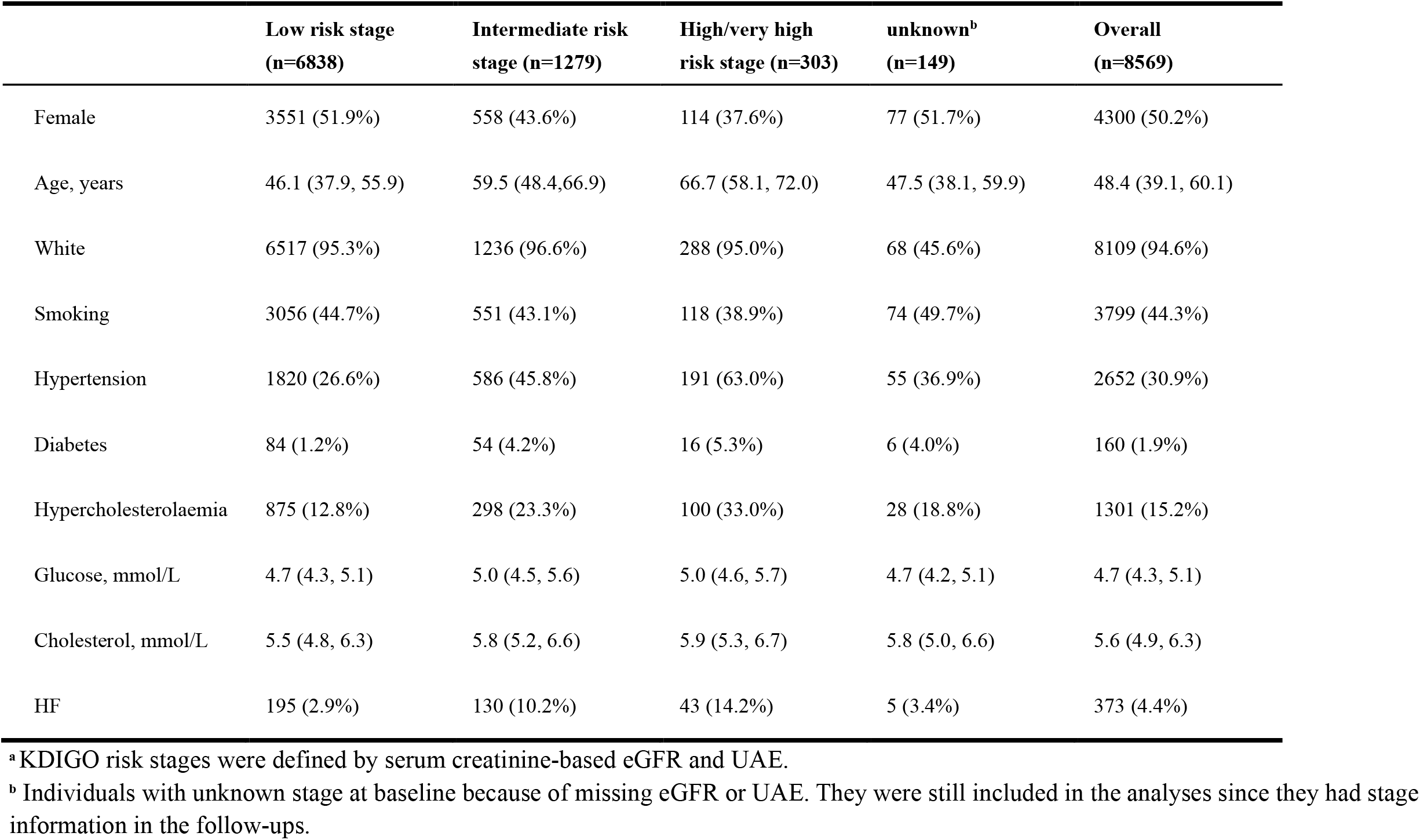
Baseline characteristics of study participants overall and by KDIGO risk stages^a^

During a median follow-up of 12.6 (interquartile range: 12.4-12.9) years, 4,973 participants (58%) completed all four visits, 961 (11%) completed three visits, 975 (11%) completed two visits, and 1,660 (19%) completed only the baseline visit. By the end of the follow-up period, 373 participants were diagnosed with new-onset HF, and 710 participants had died before developing HF.

### Transition rates

Estimated transition rates are plotted in Figure 2. With regard to the progression of kidney disease, transition rates for the two forward transitions (low risk to intermediate risk stage and intermediate risk to high or very high risk stage) were very low before age 50 (fewer than one transition per 100 person-years) and started to increase with age afterwards. The transition rate from low risk to intermediate risk stage did not differ significantly between women and men (HR: 1.08, 95% CI: 0.87-1.33); it reached ten transitions per 100 person-years at age 75 years, and then flattened afterwards. The transition rate from intermediate risk to high or very high risk stage increased exponentially with age, with women having lower rates (HR: 0.50, 95% CI: 0.34-0.76). The rate of the backward transition from intermediate risk to low risk stage decreased with age and flattened after age 50. Women had higher backward transition rates (twenty transitions per 100 person-years at age 30, which decreased to five transitions at age 50) than men (six transitions per 100 person-years at age 30, which decreased to one transition at age 50). Regarding the transitions to HF, all transition rates increased exponentially with age and were significantly lower in women than in men (HR: 0.70, 95% CI: 0.56-0.87).

**Figure 2.**
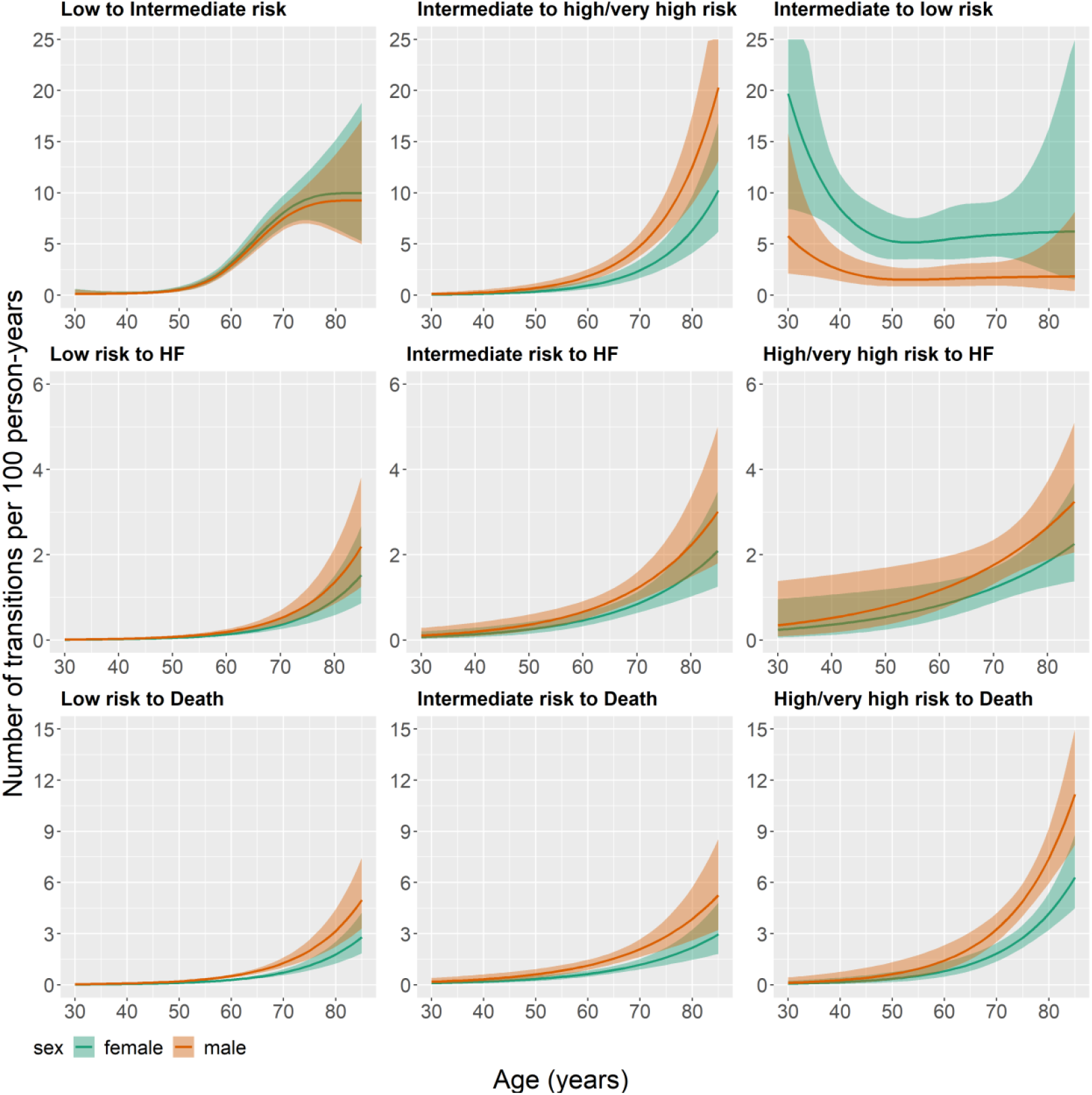
Estimated transition rates (95% CI) by age and sex. Solid lines refer to the point estimates, and shaded areas refer to 95% CI.

### Progression of KDIGO risk stage

Figure 4 depicts the predicted state frequencies for an ageing population starting in low risk stage at age 50. Over time, the proportion of subjects at low risk stage decreased dramatically, with a steeper slope after age 60. The proportion of subjects at intermediate risk stage tripled (10% to 28% for men and 10% to 30% for women) between age 60 and 70. The proportion of subjects at high or very high risk stage also increased fast (6% to 17% for men and 3% to 12% for women) but at later age between 70 and 80. The proportion of subjects who had died before developing HF increased exponentially with age and was higher in men than in women (reaching 58% at 90 years for men and 40% at 90 years for women). The proportion of subjects with new-onset HF also increased with age, reaching 24% at 90 years for men and 21% at 90 years for women.

### RRs and RDs for new-onset HF

Individuals in intermediate and high or very high risk stage had an increased risk of developing HF compared to those in the low risk stage. We found this increase in risk to be more pronounced in younger subjects (e.g., intermediate vs low risk stage: from RR=3.4 at age 60 years to RR=1.6 at 80 years; high or very high vs low risk stage: from RR=6.1 at age 60 years to RR=2 at 80 years) (Figure 3A). We did not find RRs to be different between men and women. Different from the relative scale RRs, the absolute RDs were more pronounced in older subjects. At age 80 compared with 60, RD for intermediate vs low risk stage increased from 3.3 to 6.1 transitions per 1000 person-years for women, and increased from 4.7 to 8.8 transitions per 1000 person-years for men. At age 80 compared with 60, RD for high or very high vs low risk stage increased from 6.8 to 9 per 1000 person-years for women, and increased from 9.8 to 13 per 1000 person-years for men (Figure 3B). Same trends were observed for the model adjusting for smoking, hypertension, hypercholesterolaemia and diabetes, with attenuated RRs (Figure S1). RDs were found to be more pronounced in populations with more medical histories (Figure S2 and S3).

**Figure 3.**
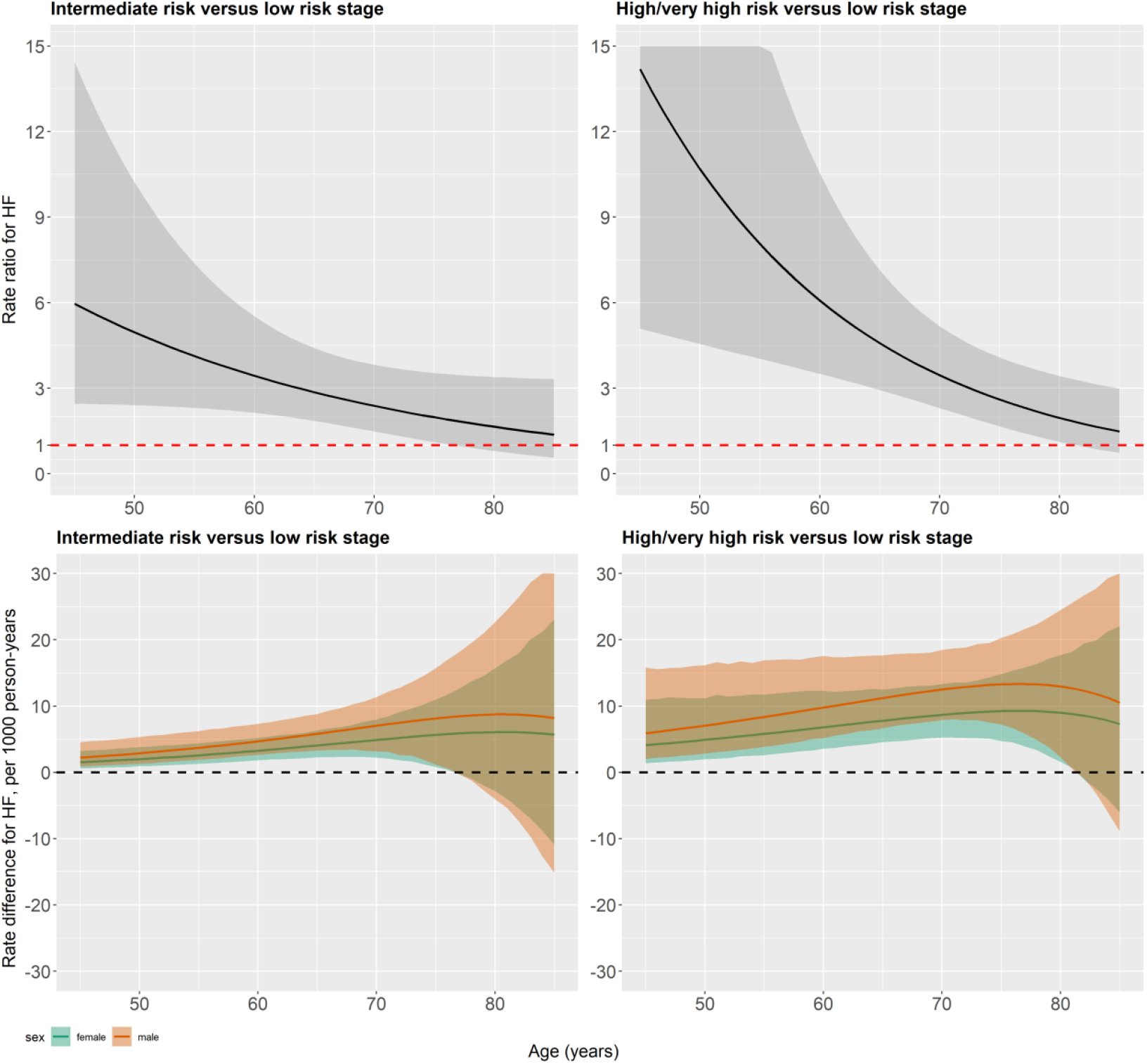
Relative Rate Ratios (with 95% CI) and absolute Rate Differences (with 95% CI) for heart failure with age. Solid lines refer to the point estimates, and shaded areas refer to 95% CI.

### Misclassification probabilities

Estimated misclassification probabilities are presented in Table 2. Individuals in low risk stage had a probability of 2.3% (95% CI: 2.0-2.8%) to be misclassified as intermediate risk stage. Individuals in intermediate risk stage had a probability of 18.4% (95% CI: 14.8-22.5%) to be misclassified as low risk stage and had a probability of 3.9% (95% CI: 3.0-4.9%) to be misclassified as high or very high risk stage. Individuals in high or very high risk stage had a probability of 15.6% (95% CI: 11.8-20.2%) to be misclassified as intermediate risk stage.

**Table 2.** Estimated misclassification probabilities

| Observed KDIGO risk stage | Actual KDIGO risk stage | Probabilities (95% CI) |
| --- | --- | --- |
| 1 | 1 | 97.7% (97.2-98.0%) |
| 2 | 1 | 2.3% (2.0-2.8%) |
| 1 | 2 | 18.4% (14.8-22.5%) |
| 2 | 2 | 77.8% (72.1-82.5%) |
| 3 | 2 | 3.9% (3.0-4.9%) |
| 2 | 3 | 15.6% (11.8-20.2%) |
| 3 | 3 | 84.4% (79.8-88.2%) |

### Model calibration

The goodness-of-fit of our model was excellent, with predicted cumulative incidence curves of HF and death very close to the observed cumulative incidence curves as estimated by the Aalen-Johansen estimator (Figure 5).

**Figure 4.**
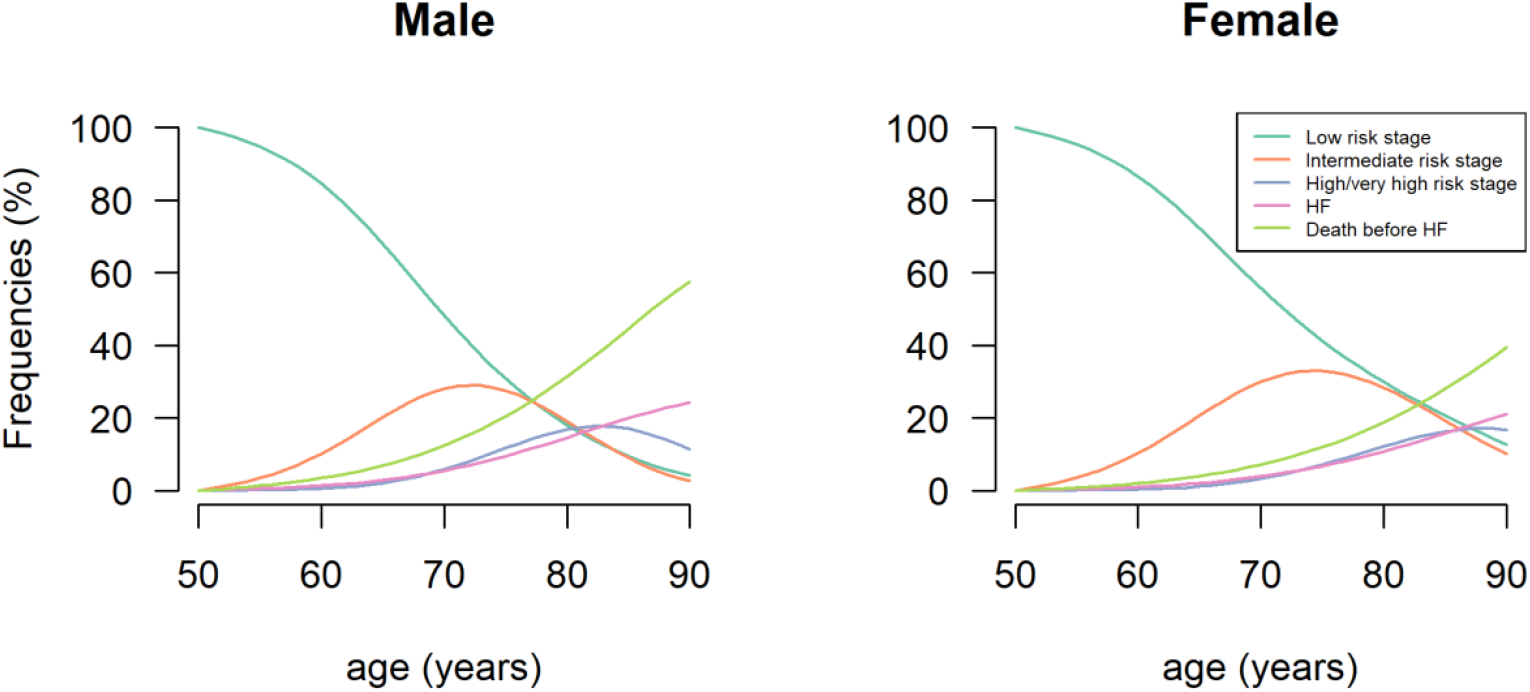
Estimated frequency of states by age and sex for an ageing population starting at KDIGO low risk stage.

**Figure 5.**
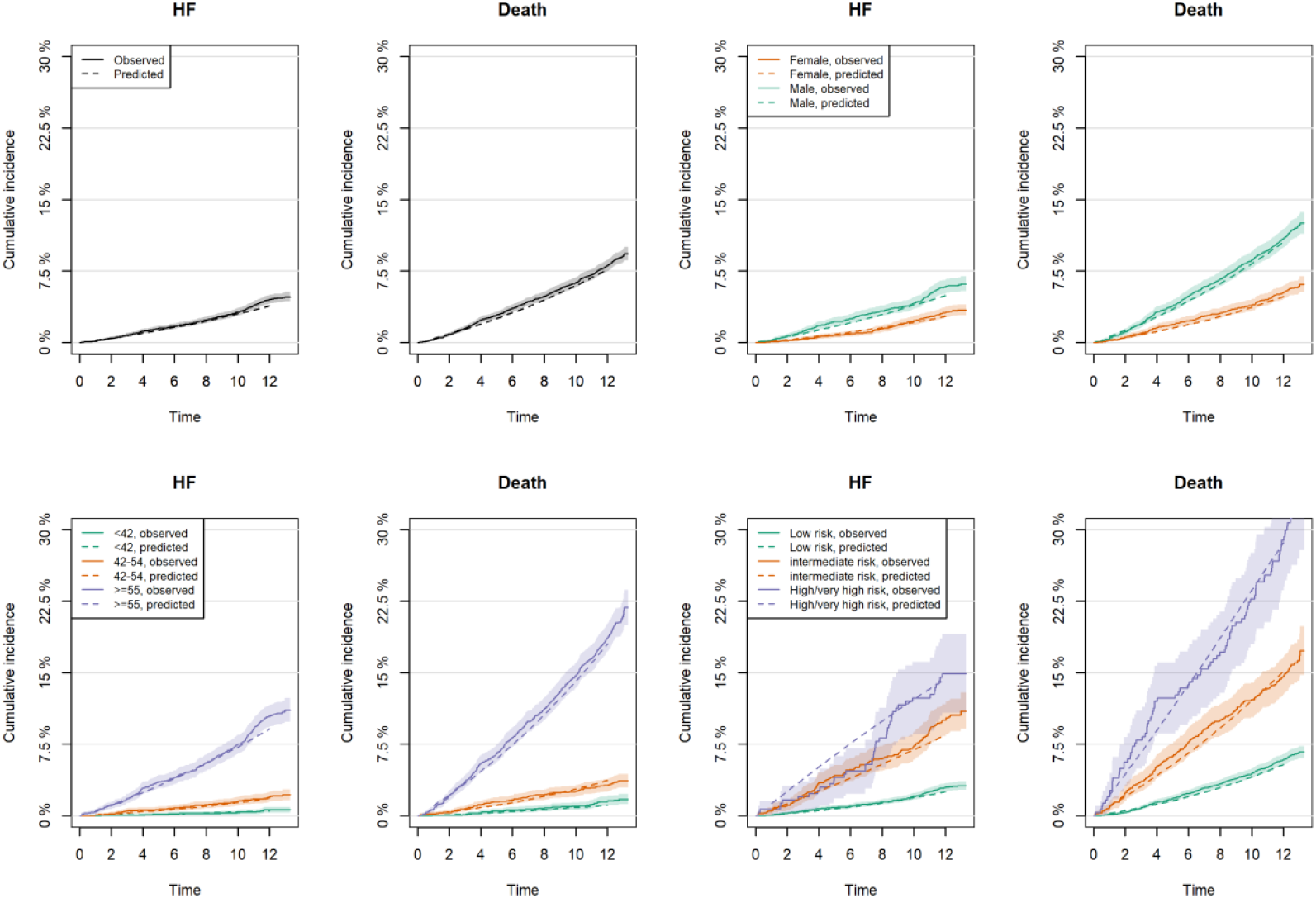
Predicted versus observed cumulative incidence curve of heart failure and death. Predicted cumulative incidence curve: dashed lines; Observed cumulative incidence curve: solid lines (point estimates) and shaded areas (95% CI). Panel A: total population; Panel B: stratification by sex; Panel C: stratification by tertiles of age at baseline; Panel D: stratification by KDIGO risk stage at baseline.

### Sensitivity analyses

Sensitivity analysis using serum cystatin C-based eGFR showed mostly consistent results (Supplemental material section 2). However, we found some differences. The transition rate from low risk to intermediate risk stage was higher in men than in women and it continued to increase after 75 years. The transition rate from intermediate risk back to low risk stage decreased after 60 years and was close to 0 after 70 years (Figure S4). The misclassification probabilities of KDIGO risk stages were lower when using cystatin C-based eGFR compared to using serum creatinine-based eGFR (Table 2 and Table S2).

## Discussion

Using repeated measurements of kidney measures from a longitudinal cohort with scheduled follow-ups, this study is one of the first to quantify the transitions of KDIGO risk stages and association with new-onset HF. We report three major findings. First, forward transition rates across KDIGO risk stages increased exponentially with age and were higher in men than in women; Younger subjects and women are more likely to make a backward transition from intermediate risk to low risk stage. Second, KDIGO risk stage progresses fast, with the progression to intermediate risk stage mostly accurring between age 60 and 70 and the progression to high/very high risk stage accurring after age 70. Third, more advanced KDIGO risk stages are associated with increased risk of new-onset HF, and the risk association attenuates with age on the relative scale but strengthens with age on the absolute scale.

### Comparison with previous studies

Most previous studies^23,24^ investigated the risk of developing HF based on a single snapshot of a subject’s kidney function at baseline. Our investigation extended these previous studies by taking advantage of a longitudinal cohort with scheduled follow-ups and modelling a subject’s entire disease course from the initial kidney function risk stage at baseline to the development of HF. We found the forward transition rates from low risk to intermediate risk stage and from intermediate risk to high or very high risk stage to rapidly increase with age, suggesting that even a small reduction in these transition rates in middle-aged subjects by an intervention may result in an important reduction in the incidence of new-onset HF. The backward transition rate from intermediate risk to low risk stage was high at younger age, to stabilize at low levels from 50 years and on. This observation was not changed by using the serum cystatin-based eGFR, suggesting that kidney dysfunction is more likely to reverse in younger subjects.

Previous population-based studies yielded inconsistent results about sex differences in the change of kidney function with age^25–29^. Most of these studies focused on eGFR alone, did not consider the recovery of kidney function, and did not account for loss of follow-up because of death. We found that woman was protective in the evolution of kidney function risk stages with lower forward transition rates and higher backward transition rates compared with men (Figure 2 and Figure S1). This may be explained by the protective effects of oestrogens in women and/or the damaging effect of testosterone, as well as unhealthier lifestyles in men^5,23,30^.

The transition from low risk to intermediate risk stage deserves special attention because this transition rate did not increase exponentially with age. This rate increased rapidly from 0 at 50 to ten transitions per 100 person-years at 75 years, and plateaued from 75 years. However, we are not confident about this finding because of the wide confidence intervals after 75 years (Figure 2). We hypothesized this was most likely explained by a progressive decline of muscle mass with age that may lead to bias in serum creatinine-based eGFR^16^. The sensitivity analysis using serum cystatin C-based eGFR confirmed our hypothesis by showing that the transition rate from low risk to intermediate risk stage did exponentially increase with age (Figure S1).

In line with previous studies^28^, we found that, although men were more likely to develop HF, they were also more likely to die in general, irrespective of age and KDIGO risk stage. This may partly explain the comparable lifetime risk of HF between the sexes as shown in the Framingham Heart Study^31^.

Previous studies, including the CKD-PC observed weaker relative risks but stronger absolute risks of renal dysfunction in older subjects for all-cause and cardiovascular mortality^4,32,33^. Our study extended these findings to the development of HF. Compared with previous studies, our study conducted a more robust analysis by accounting for both dynamic change of kidney function with age and potential misclassification of kidney function risk stage. The decreasing relative risk with age could be explained by the higher baseline risk of new-onset HF for older subjects due to other factors than renal dysfunction. Although the decreasing relative risk with age may support an age-dependent staging for CKD, the increasing absolute risk with age defends the current age-independent CKD staging system in the guidelines. The latter suggests that more HF cases would be prevented in the older population if effective preventive or treatment strategies were considered. However, further studies are needed to investigate whether those strategies are equally effective and cost-effective across the age spectrum.

### Strength and limitations of the study

A strength of our study is that we accounted for the potential misclassification of kidney function risk stage in our model. Previous studies showed serum creatinine measurements that are used to calculate eGFR may vary due to diet and other environmental factors^34^; UAE may also increase after strenuous exercise^35^. Ignoring the potential misclassification of the risk stage due to the high variability of serum creatinine and UAE can lead to false transitions between risk stages. Our study indicates that an actual intermediate risk stage is frequently misclassified as low risk stage (18.4%, 95% CI: 14.8-22.5%) and an actual high or very high risk stage is frequently misclassified as intermediate risk stage (15.6%, 95% CI: 11.8-20.2%). Interestingly, we found the misclassification probabilities were lower for serum cystatin C-based eGFR, suggesting that serum cystatin C-based might be more accurate in staging CKD compared to serum creatinine-based eGFR. Another strength is that our multi-state model took into account interval-censored data. In our study, KDIGO risk stage was not observed continuously with time, but was periodically assessed at clinic visits. Therefore, the stage-to-stage transition was only known to have occurred within a bounded time interval, usually between visits. Assuming that the assessment times are the exact times of transition can provide misleading results^36^. Other strengths of this study include the use of repeated measurements of kidney function for both eGFR and UAE and a well-validated endpoint.

Our study has some limitations. First, the PREVEND cohort was enriched for increased UAE, meaning that this cohort is not a random sample from the general population. However, the association between KDIGO risk stage and new-onset HF should not be altered since UAE was used to define the risk stage^37^. Second, subjects in PREVEND were predominantly Caucasian, limiting the generalization to other populations. Finally, the Markov assumption implies that the estimated transition rates were assumed to depend only on the current risk stage and the age of the subject at that time point and not on other aspects of a subject’s disease history.

## Conclusions

Kidney disease progresses fast in the middle-aged population. A more advanced KDIGO status is associated with increased risk of new-onset HF. Together, These findings indicate that slowing the progression of KDIGO risk stages may result in an important reduction in the incidence of new-onset heart failure in this population of middle-aged subjects.

## Data Availability

All data produced in the present study are available upon reasonable request to the authors.

