## Supplementary material for "Transitions across Kidney Disease Stages and Association with New-onset Heart Failure: a Population-Based study of 8569 Individuals and 12.6 Years of Follow-up"

**Authors:** Yuntao Chen, Stephan JL Bakker, Navin Suthahar, Laura M.G. Meems, [Dion Groothof,](https://www.sciencedirect.com/science/article/pii/S0025619619309425?via%3Dihub#!) Dirk J. van Veldhuisen, Wiek H. van Gilst, Rudolf A de Boer, Ron T Gansevoort, Gerton Lunter, Hans L. Hillege, Douwe Postmus

**Table of Contents**

**Section 1:** Adjusted rate ratios and rate differences based on model further adjusting for smoking, hypertension, hypercholesterolaemia and diabetes. ----------------------------Page 2-4

**Section 2**: we reproduced all the results in the main text (tables and figures) by replacing serum creatinine-based eGFR with cystatin C-based eGFR. ------------------------------------Page 5-10

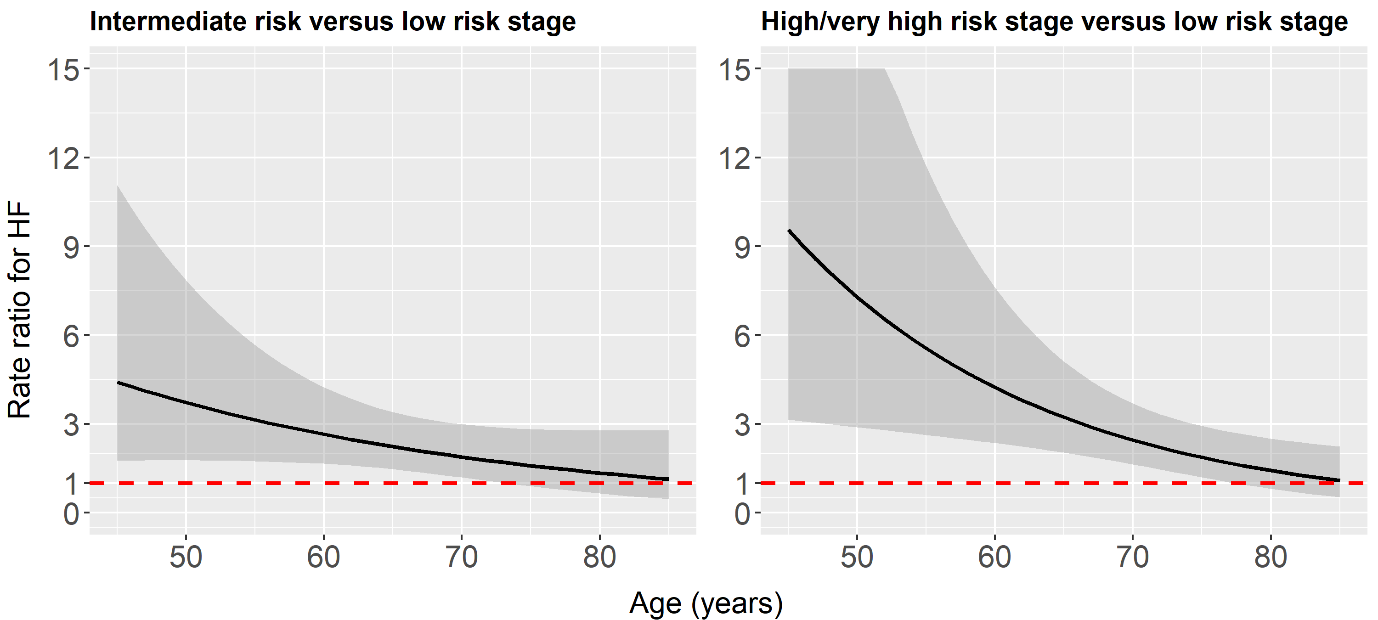

Figure S1. Relative rate ratios (with 95% CI) for heart failure with age. Solid lines refer to the point estimates, and shaded areas refer to 95% CI. This model was adjusted for smoking, hypertension, hypercholesterolaemia and diabetes.

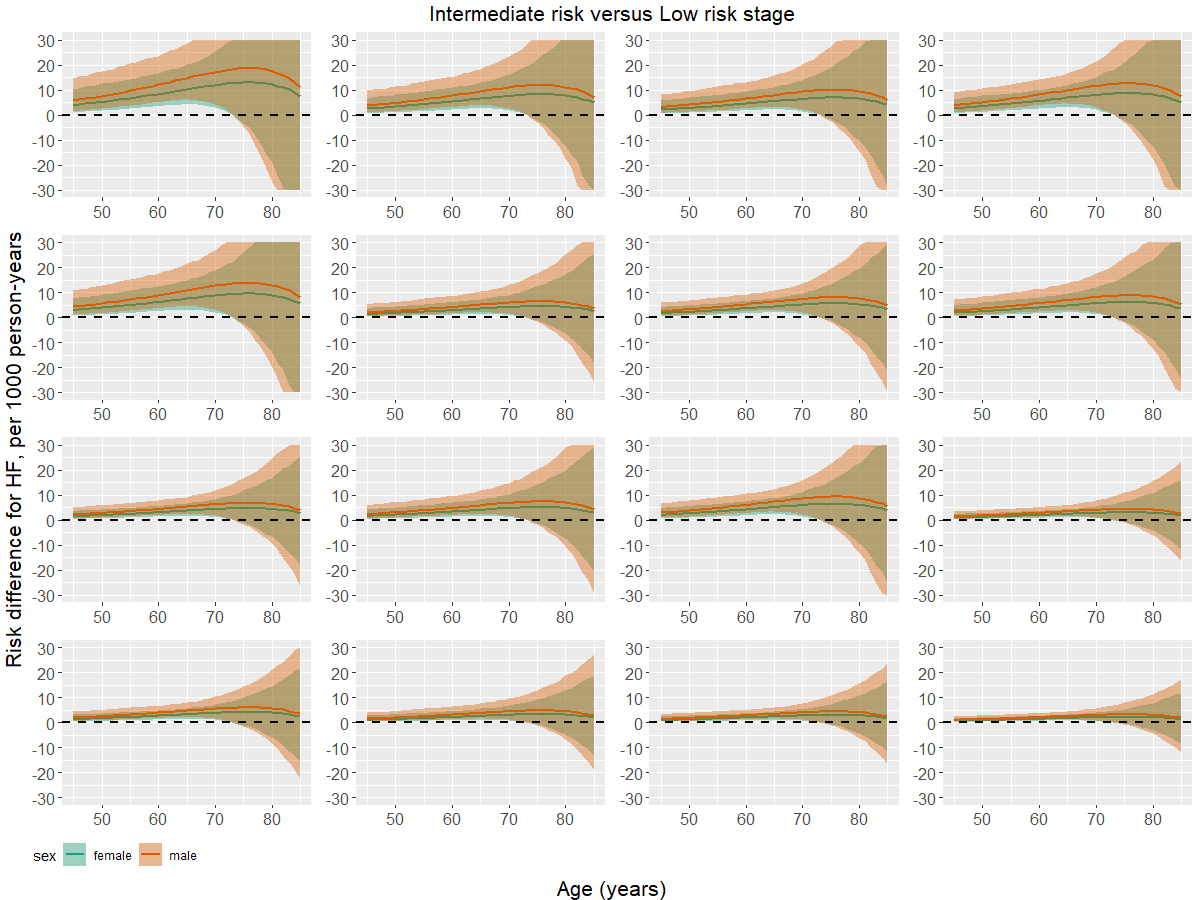

Figure S2. Absolute rate differences (95% CI) for heart failure with age. Solid lines refer to the point estimates, and shaded areas refer to 95% CI. This model was adjusted for smoking, hypertension, hypercholesterolaemia and diabetes. 16 panels were plotted depending on covariate values (smoking, hypertension, hypercholesterolaemia and diabetes). Panel 1: population with all four medical histories; Panel 2-5 (ordered by row): populations with three medical histories; Panel 6-11: populations with two medical histories; Panel 12-15: populations with one medical histories; Panel 16: population without those four medical histories.

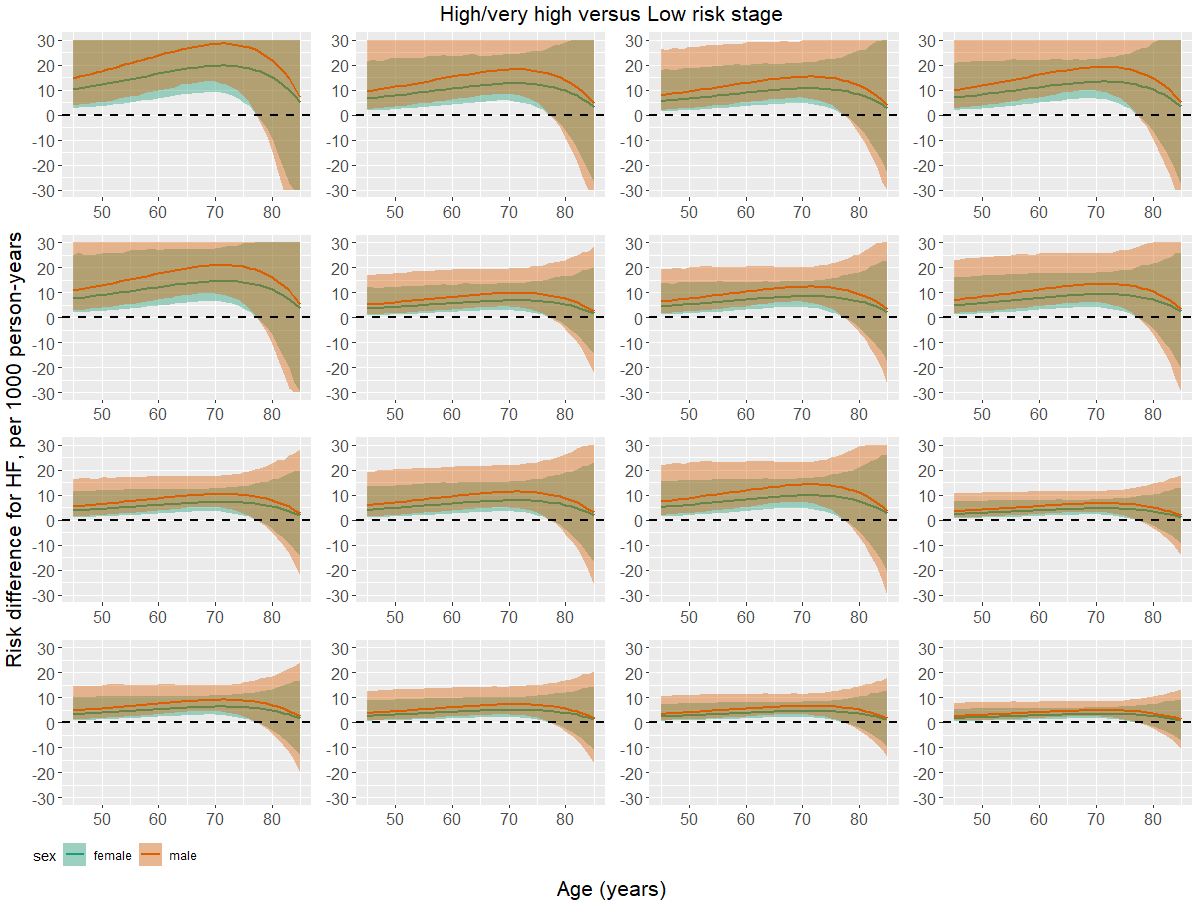

Figure S3. Absolute rate differences (95% CI) for heart failure with age. Solid lines refer to the point estimates, and shaded areas refer to 95% CI. This model was adjusted for smoking, hypertension, hypercholesterolaemia and diabetes. Panel order was same as Figure S2, with absolute rate differences for HF (High/very high risk stage versus Low risk stage).

Table S1. Baseline characteristics of study participants overall and by KDIGO risk stages^a^

|  | **Low risk stage  (n=6533)** | **Intermediate risk stage (n=1147)** | **High/very high risk stage (n=318)** | **unknown^b^ (n=571)** | **Overall (n=8569)** |
| --- | --- | --- | --- | --- | --- |
| Female | 3457 (52.9%) | 470 (41.0%) | 105 (33.0%) | 268 (46.9%) | 4300 (50.2%) |
| Age, years | 46.2 (37.9, 55.9) | 59.0 (47.8,66.9) | 66.8 (59.2, 72.0) | 50.0 (39.1, 61.1) | 48.4 (39.1, 60.1) |
| White | 6182 (94.6%) | 1097 (95.6%) | 299 (94.0%) | 531 (93.0%) | 8109 (94.6%) |
| Smoking | 2873 (44.0%) | 540 (47.1%) | 136 (42.8%) | 250 (43.8%) | 3799 (44.3%) |
| Hypertension | 1748 (26.8%) | 502 (43.8%) | 199 (62.6%) | 203 (35.6%) | 2652 (30.9%) |
| Diabetes | 74 (1.1%) | 49 (4.3%) | 19 (6.0%) | 18 (3.2%) | 160 (1.9%) |
| Hypercholesterolaemia | 847 (13.0%) | 255 (22.2%) | 101 (31.8%) | 98 (17.2%) | 1301 (15.2%) |
| Glucose, mmol/L | 4.6 (4.3, 5.0) | 5.0 (4.5, 5.5) | 5.0 (4.6, 5.8) | 4.9 (4.6, 5.4) | 4.7 (4.3, 5.1) |
| Cholesterol, mmol/L | 5.5 (4.8, 6.3) | 5.8 (5.2, 6.6) | 5.9 (5.2, 6.7) | 5.7 (5.0, 6.4) | 5.6 (4.9, 6.3) |
| HF | 176 (2.7%) | 121 (10.5%) | 51 (16.0%) | 25 (4.4%) | 373 (4.4%) |

**^a^** KDIGO risk stage was defined by serum cystatin C-based eGFR and UAE.

**^b^** Individuals with unknown stage at baseline because of missing eGFR or UAE. They were still included in the analyses since they had stage information in the follow-ups.

Table S2. Estimated misclassification probabilities

| **Observed KDIGO risk stage** | **Actual KDIGO risk stage** | **Probabilities (95% CI)** |
| --- | --- | --- |
| 1 | 1 | 98.6% (98.2-98.9%) |
| 2 | 1 | 1.4% (1.1-1.8%) |
| 1 | 2 | 12.9% (10.4-15.9%) |
| 2 | 2 | 84.7% (78.9-89.1%) |
| 3 | 2 | 2.4% (1.7-3.4%) |
| 2 | 3 | 8.4% (5.9-11.8%) |
| 3 | 3 | 91.6% (88.2-94.1%) |

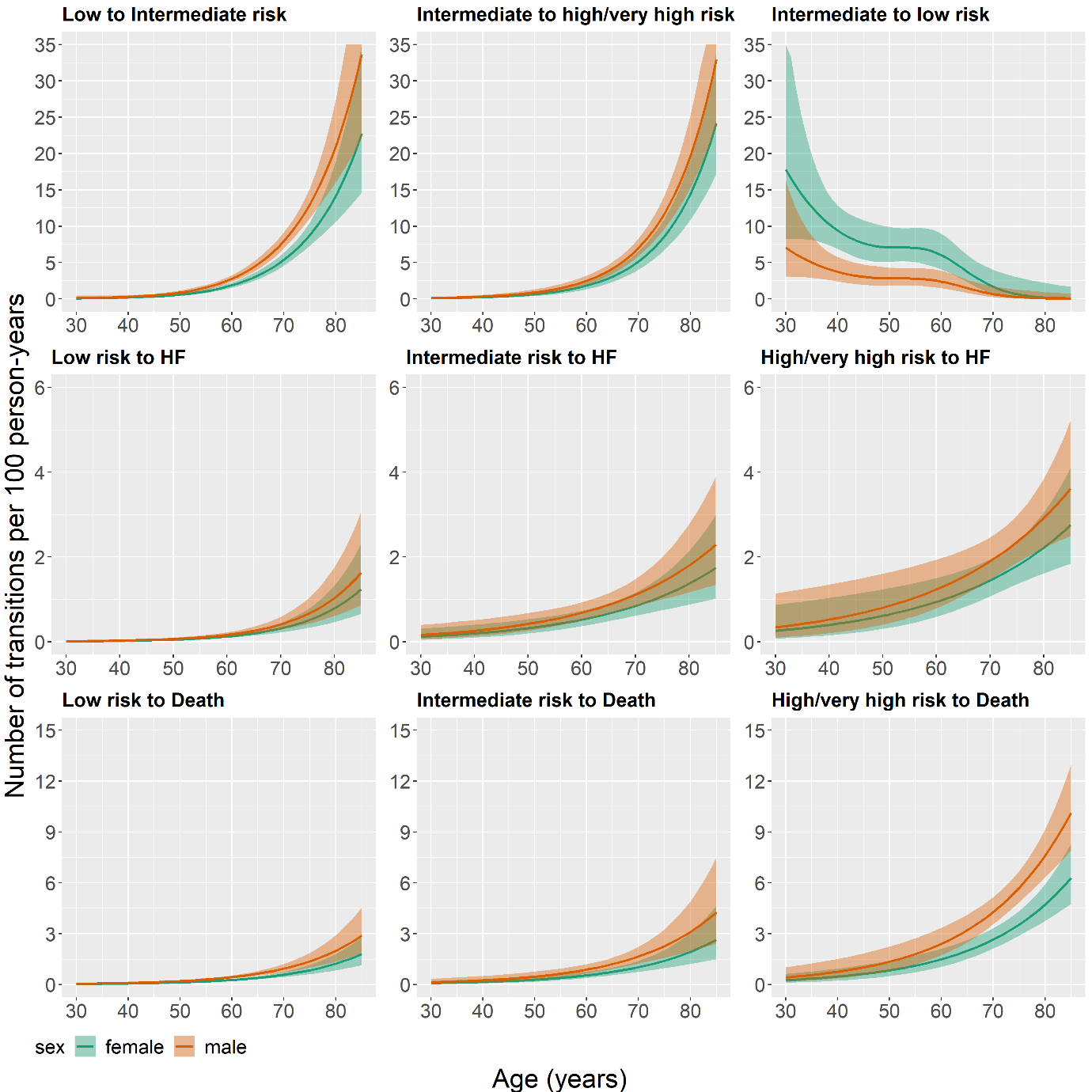

Figure S4. Estimated transition rates (95% CI) by age and sex. Solid lines refer to the point estimates, and shaded areas refer to 95% CI.

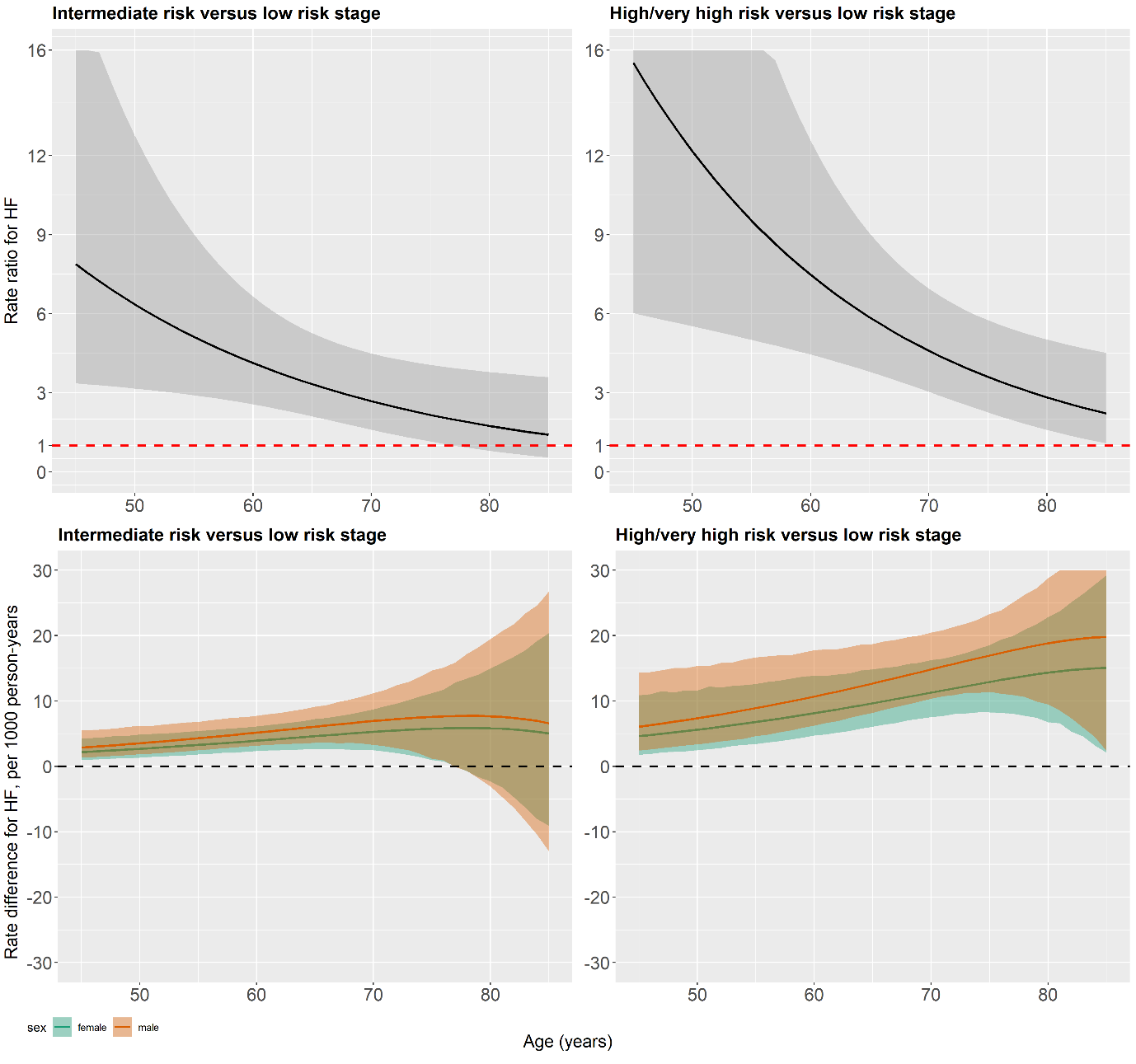

Figure S5. Relative rate ratios (with 95% CI) and absolute rate differences (95% CI) for heart failure with age. Solid lines refer to the point estimates, and shaded areas refer to 95% CI.

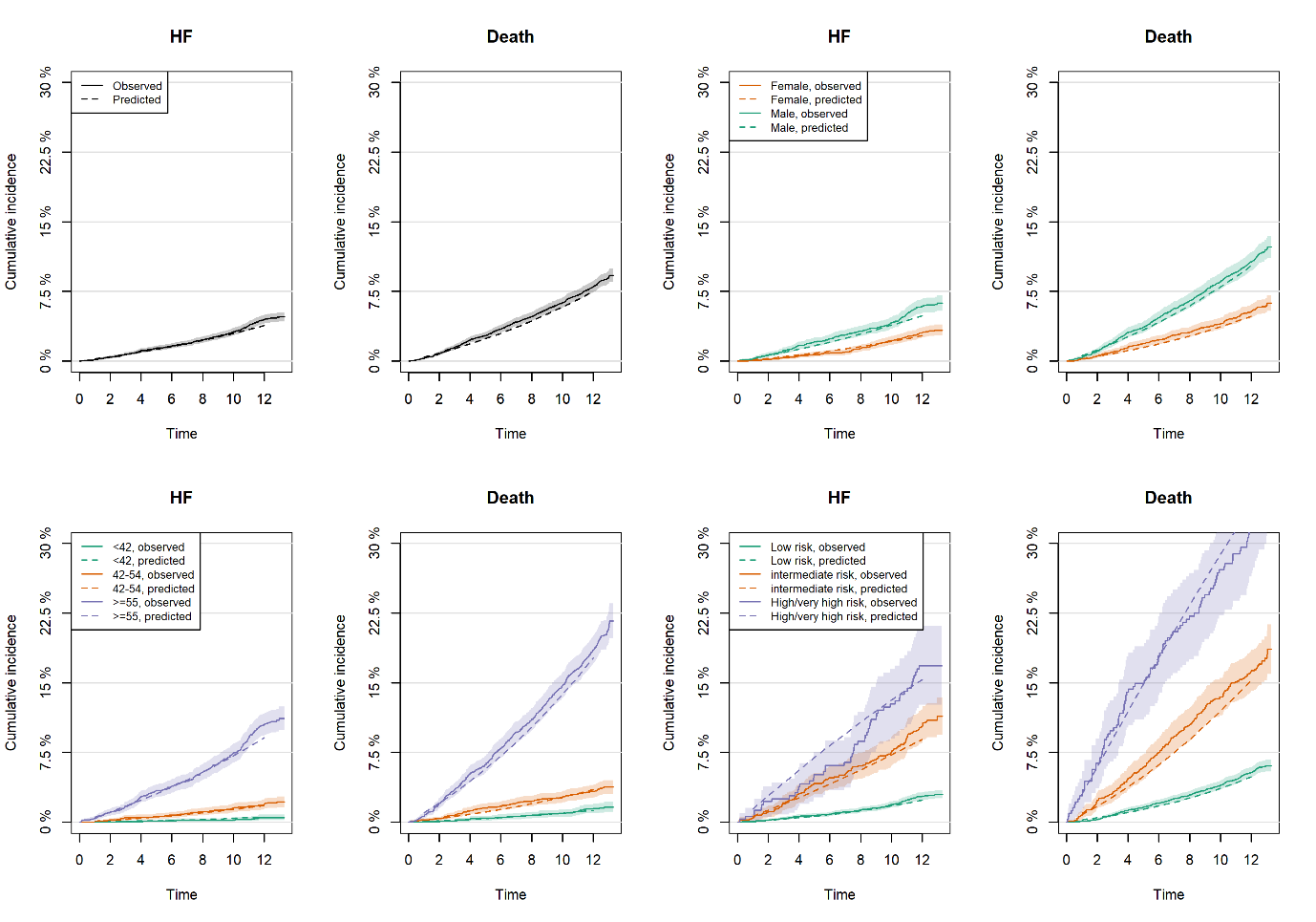

Figure S6. Predicted versus observed cumulative incidence curve of HF and death. Predicted cumulative incidence curve: dashed lines; Observed cumulative incidence curve: solid lines (point estimates) and shaded areas (95% CI). Panel A: total population; Panel B: stratification by sex; Panel C: stratification by tertiles of age at baseline; Panel D: stratification by KDIGO risk stage at baseline.

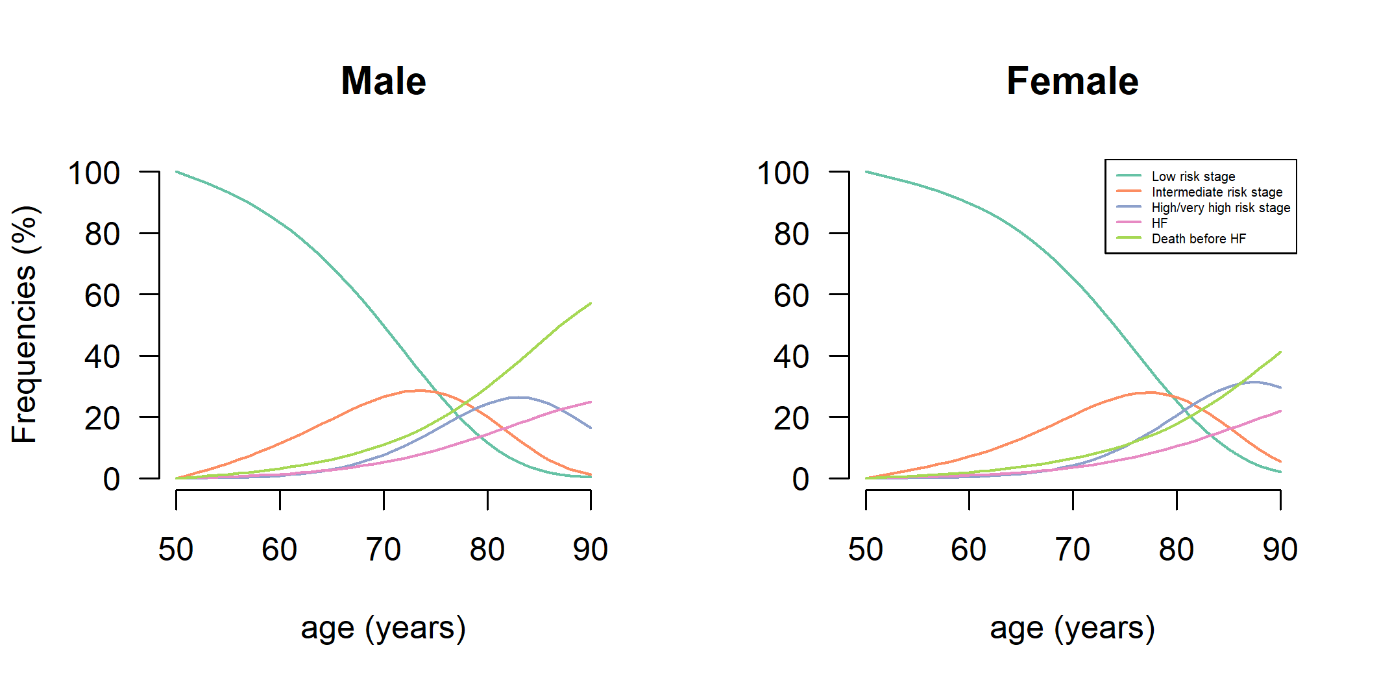
Figure S7. Estimated frequency of states by age and sex for an ageing population starting at KDIGO low risk stage at age 50.
